# Monitoring the propagation of SARS CoV2 variants by tracking identified mutation in wastewater using specific RT-qPCR

**DOI:** 10.1101/2021.03.10.21253291

**Authors:** S Wurtzer, P Waldman, M Levert, JM Mouchel, O Gorgé, M Boni, Y Maday, OBEPINE consortium, V Marechal, L Moulin

**Affiliations:** Eau de Paris, Département de Recherche, Développement et Qualité de l’Eau, 33 avenue Jean Jaurès, F-94200 Ivry sur Seine, France; Sorbonne Université, CNRS, Université de Paris, Laboratoire Jacques-Louis Lions (LJLL), F-75005 Paris, France; Sorbonne Université, CNRS, EPHE, UMR 7619 Metis, e-LTER Zone Atelier Seine, F-75005 Paris, France; Institut de Recherche Biomédicale des Armées, Division Défense NRBC, Microbiologie et Maladies Infectieuses, 1 place Valérie André, F-91220 Brétigny sur Orge, France; Institut de Recherche Biomédicale des Armées, Direction scientifique et technique, moyens expérimentaux partagés, 1 place Valérie André, F-91220 Brétigny sur Orge, France; Sorbonne Université, INSERM, Centre de Recherche Saint-Antoine, F-75012, Paris, France

**Keywords:** SARS-CoV-2, COVID-19, variant, mutation, wastewater, epidemiology

## Abstract

Since the end of 2020, the COVID-19 pandemic has experienced a major turning point with the appearance and rapid spread of new variants, causing a significant increase in the number of new cases requiring hospitalization. These so-called UK, South African or Brazilian variants are characterized by combinations of mutations which allow them to be distinguished from the variants which have circulated since the start of the epidemic. The impact of these variants on the functioning of healthcare systems requires monitoring the spread of these variants, which are more contagious, more lethal and may reinfect people who are already immune to a natural infection or to a vaccination. Monitoring the viral genome in wastewater has shown great value in early detection of the dynamics of virus spreading in populations.

The sequencing of viral genomes is used in humans, but its application and interpretation on wastewater matrices are much more complex due to the diversity of circulating strains. Also this study demonstrates the possibility of following certain mutations found in these new variants by targeted RT-qPCR. This study is the first carried out in France demonstrating the spreading dynamics of the 69-70 deletion in the Spike protein of SARS-CoV-2.

## Introduction

The first case of coronavirus disease was identified in December 2019, although recent publications (including wastewater epidemiology approach) suggested evidences of an earlier virus circulation in some area ((Deslandes et al. 2020; Carrat et al. 2021; La Rosa et al. 2021). Since the start of what has become a worldwide pandemic more than one hundred million cases were diagnosed and 2,2 million people died up to January 2021. COVID-19 results from infection by SARS-CoV-2, an RNA virus belonging to the Coronaviridae family. Like many other single stranded-RNA viruses (ssRNA), SARS-CoV-2 genome contains an RNA-dependent RNA polymerase (RdRp) which is implicated in subgenomic mRNA synthesis for producing the different viral proteins and in pregenomic mRNA synthesis, which are carried away during the virus budding. Despite having proof-reading capability for stabilizing the 29.9 kbases long genome, the mutation rate observed in the SARS-CoV-2 genome is comparable to other ssRNA viruses that lack such capability (Y.-Z. Zhang et Holmes 2020). Indeed SARS-CoV-2 variability may occur from a combination between uncorrected mutations linked to genome replication, recombination and RNA editing by host deaminases (Giorgio et al. 2020). In this context, SARS-CoV-2 genomes have evolved from the ancestral Wuhan-1 strain to diverse SARS-CoV-2 lineages under the influence of various selective pressures.

SARS-CoV-2 genome sequencing in human and animal allowed to monitor the geographic spread of the virus since its emergence. If variants are regularly described, the recent identification of variants with an increased infectivity, an expanded severity, a longer excretion time (Kissler et al. 2021) and/or a putative ability to partially escape natural or vaccination-induced immunity deserved a particular attention (Francisco et al. 2021; Li et al. 2020; Wibmer et al. 2021; Wang et al., s. d.; Cele et al. 2021). These notably include B.1.1.7 (or 501Y.V1), firstly isolated in the UK (Rambaut et al. 2020), B.1.351 (or 501Y.V2) identified in South Africa (Tegally, Wilkinson, Giovanetti, et al. 2020), B.1.1.28 or P1 (or 501Y.V3) detected in Brazil (Francisco et al. 2021), and more recently the strain named CAL.20C which emerged in Southern California (W. Zhang et al. 2021). Due to their increased infectivity and longer excretion period, these variants spread locally to become the predominant strain in a short period of time (Volz et al. 2021; Walker et al. 2021; Tegally, Wilkinson, Lessells, et al. 2020). These variants are characterized by mutations located in the Spike protein, especially in the receptor binding domain (RBD) that allows the interaction with the cellular receptor ACE2. The deletion ΔHV 69-70 in the Spike protein is out of RBD domain and was notably found in B.1.1.7 among recent circulating variants. This deletion was previously identified among different lineages, without any evidence of increased transmissibility (van Dorp et al. 2020). Deletion of these residues might allosterically change S1 conformation (Wrapp et al. 2020). The RBD domain is the main target of neutralizing antibodies produced during natural infection. Among mutations reported in the RBD, the mutation N501Y that is common to UK, South African and Brazilian variants, is associated with an increased transmission by enhancing binding to the ACE2 receptor (Wrapp et al. 2020; Zhao et al. 2021; Villoutreix et al. 2021), while the mutation E484K that is found in B.1.351 and B.1.1.28, is associated with a less efficient sero-neutralisation by antibodies raised against the wild-type spike protein. However, the recent finding of E484K mutation in some UK variant strains is a remarkable example of evolutionary convergence (Wise 2021).

The monitoring of SARS-CoV-2 genomes in raw wastewater was successfully set up for estimating the dynamics of viral pandemic in population linked to a wastewater network. Since both symptomatic and asymptomatic individuals are thought to contribute to wastewater inputs, the analysis of samples from sewage plants provide a more comprehensive picture of SARS-CoV-2 genomic diversity circulating in a population than clinical testing and sequencing alone. Some studies described the genomic analysis of wastewater samples. The presence of the more prevalent lineages can be suspected in wastewater samples using a combination of single nucleotide variation analysis, which can be compared to the presence of variant in patients (Fontenele et al. 2021; Crits-Christoph et al. 2021; Jahn et al. 2021; Lara et al. 2020; Martin et al. 2020; Nemudryi et al. 2020). To our knowledge, no study report the tracking of those novel variants in France, especially lineages B.1.1.7 and B.1.351, which were firstly identified the 25th and the 31st of December 2020 respectively (Bal et al. 2021). In order to evaluate the spread of these variants in France, 2 flash surveys based on intensive sequencing were done to date and showed the progression of UK variants. The prevalence of UK variants was the 2.5% by January 7, 2021 and 9,4% by January 21, 2021 in Greater Paris, France.

In the present study, we could detect mutations related to SARS-CoV-2 variants in raw wastewater samples in Greater Paris area, France. Next, we determined the dynamics of the proportion of the del69-70 mutation, probably related to the UK variant B.1.1.7 from January to mid February 2021 by using RT-qPCR targeting variant-specific polymorphism.

## Material and methods

### Design of RT-qPCR specific of the deletion 69-70

Based on sequences published in GISAID database, new couples of oligonucleotides were designed using AlleleID 9.0 software. Specificity and sensitivity were evaluated on viral RNA kindly provided by the French Armed Forces Biomedical Research Institute. The best primers and probe are listed in table 1. The amplification was done using Fast virus 1-step Master mix 4x (Lifetechnologies).

**Table 1.**
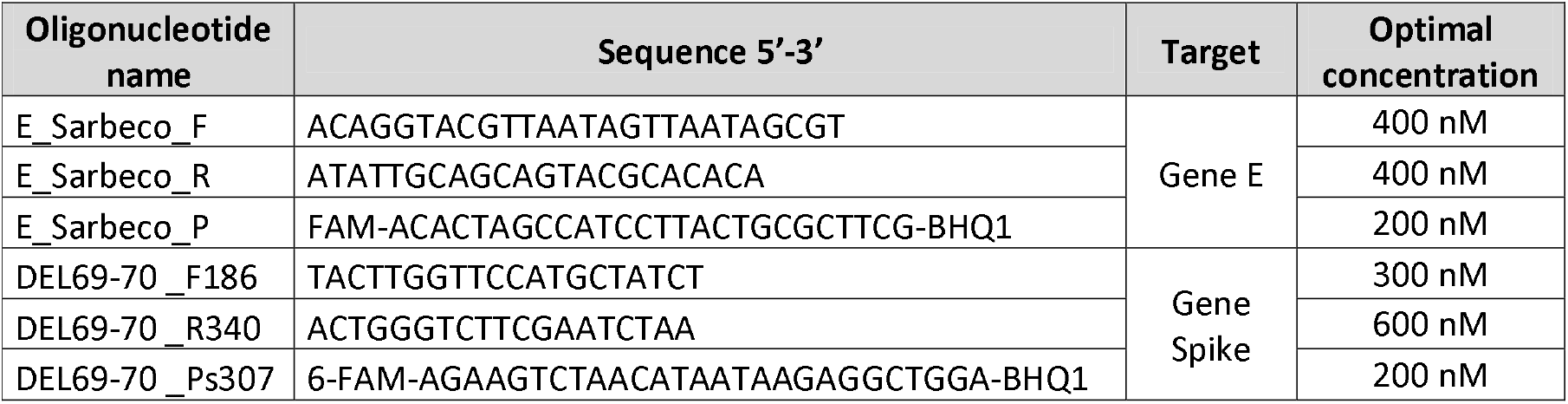
Primers and double dye probes for the specific amplification of the deletion HV69-70 and gene E in SARS-CoV-2 genome

**Table 2.** Amplification results of two RT-qPCR (E-Sarbeco and Del69-70) on three different viral strains after 45 amplification cycles.

|  | gene E (E-Sarbeco) | Del69-70 |
| --- | --- | --- |
| 2020 Wuhan-1 | + | - |
| 2021 UK strain | + | + |
| 2021 SA strain | + | - |

### Collection of raw wastewater, RNA isolation and genome detection

Raw wastewaters were collected at the inlet of five wastewater treatment plants (WWTP) in Greater Paris, France (see Wurtzer et al. 2020 for WWTP description). Briefly, 11 millilitres of 24h-samples were concentrated by ultracentrifugation and total RNA was extracted using a QIASymphony instrument (QIAGEN) as previously described by Wurtzer and colleagues (Wurtzer, Marechal, et al. 2020). Detection of SARS-CoV-2 was done by RT-qPCR using E-Sarbeco primers and probe (Corman et al. 2020) and del69-70 using new designed oligonucleotides. A relative quantification method was performed on QuantStudio 5 thermal cycler (Life technologies).

## Results

### Sensitivity and specificity of del69-70 RT-qPCR

For evaluating the specificity of del69-70 oligonucleotide set, viral RNA from Wuhan-1 strain, UK variant and South African variant were amplified and results compared to E-Sarbeco amplification.

The sensitivity of del69-70 RT-qPCR was evaluated using dilution of Wuhan-1 and UK variant RNA isolated from patients. The results presented in figure 1 showed a similar sensitivity and PCR efficacy of both E-Sarbeco and del69-70 RT-qPCR on UK variant (PCR efficiencies 89.8% and 84.0% respectively). Strains harboring wild type polymorphism were not amplified using del69-70 RT-qPCR. The initial viral load was different between both patients, which explains the difference between the data sets marked black and red in figure 1.

**Figure 1.**
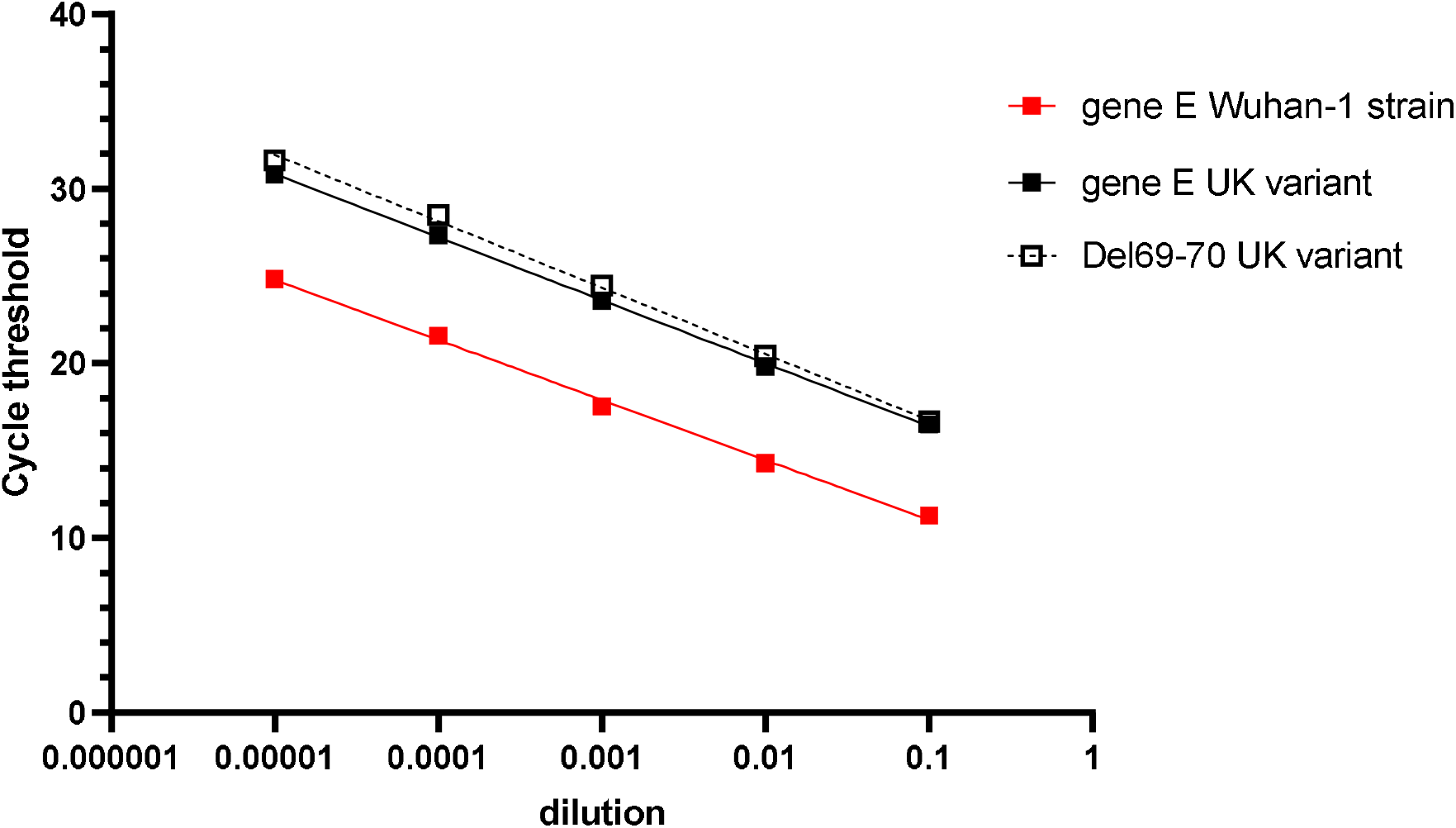
Comparison of performance for detecting UK variant using gene E-or del69-70-based amplification.

### Screening of del69-70 in SARS-CoV-2 genome from raw wastewater

Since March 2020, raw wastewaters are monitored twice a week by the Observatoire Epidémiologique dans les Eaux Usées (OBEPINE), the French sentinel network for SARS-CoV-2 in wastewater. Since the end of December 2020, patients infected with UK variant were identified in France. The deletion 69-70 was simultaneously monitored with E gene from viral genome. The relative quantification of del69-70 containing genomes (red open circles) vs wild-type genomes was performed in the 5 WWTP followed in Greater Paris (figure 2). The red curve shows the evolution of the median value. SARS-CoV-2 total concentration in wastewater (blue data points) exhibited a slight increase when del69-70-containing genomes stabilized around 30% of the circulating strains.

**Figure 2.**
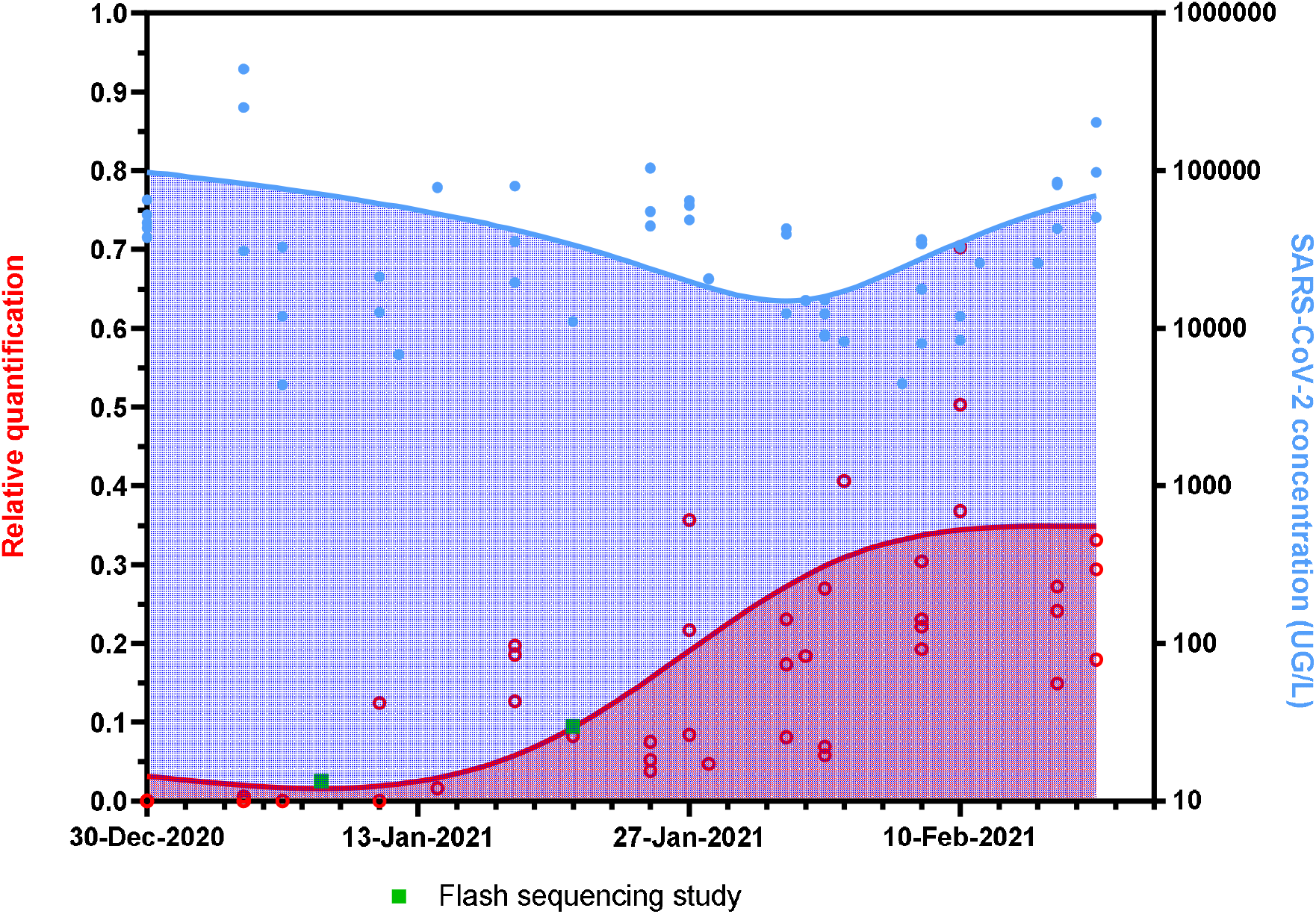
Relative quantification of del69-70 mutation in Greater Paris area wastewater samples.

Using the relative proportion of the del69-70 mutation, the absolute quantity of variant and other variants in wastewater could be calculated. The figure 3 shows the rise of SARS-CoV-2 variants carrying the deletion 69-70 from the end of 2020. Before December 15^th^, 2020, variants carrying the del69-70 could not be detected.

**Figure 3.**
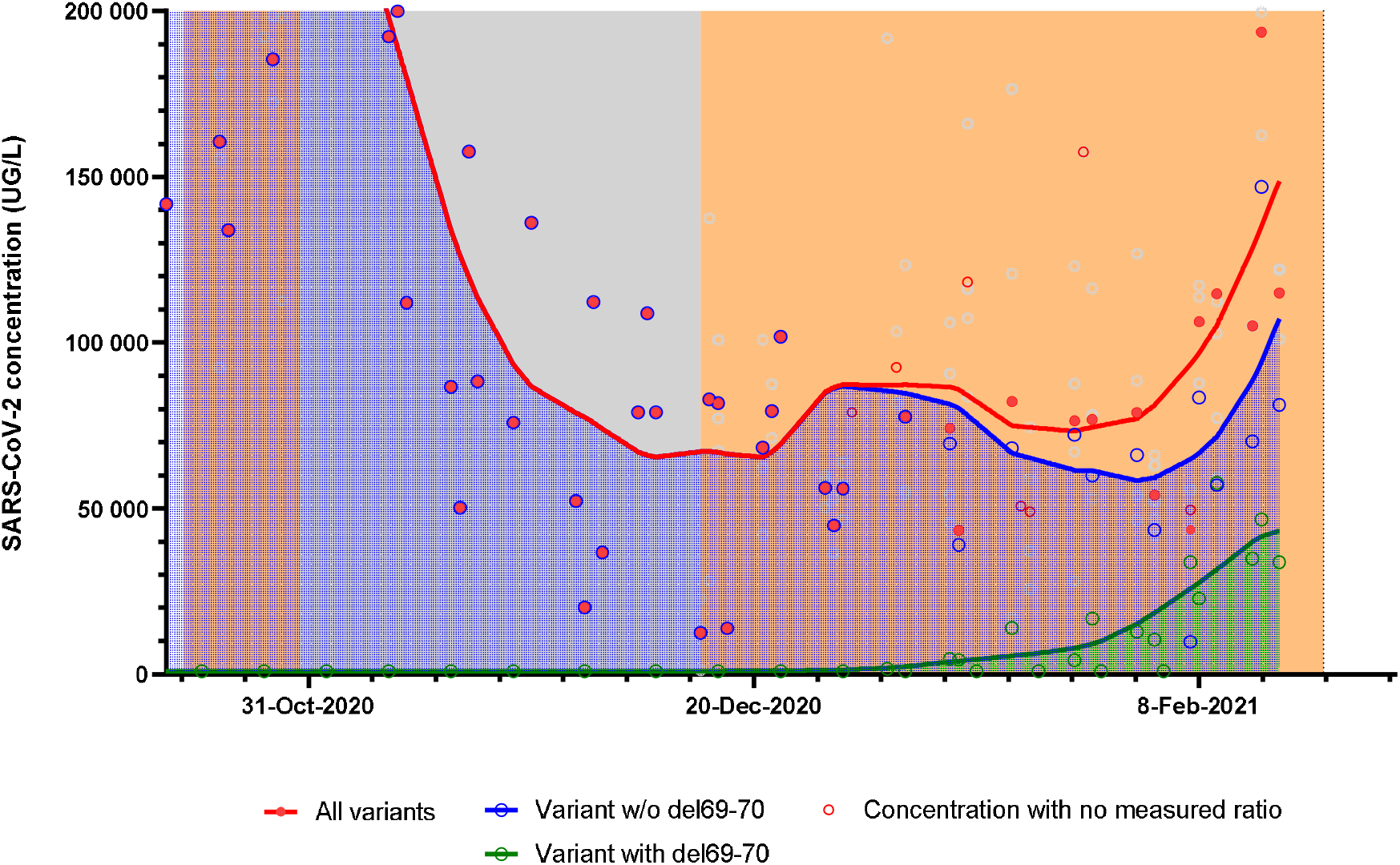
Evolution of del69-70 carrying variants and other variants in Greater Paris area wastewater. The orange zone means the curfew period and the grey zone corresponds to the lock-down period.

## Discussion

The present study is the first temporal analysis investigating the proportion of variants harbouring del69-70 in Greater Paris WWTP. Although this mutation cannot be considered as specific from the B.1.1.7 variant, it is likely that the recent predominance of this mutation in viral strains collected from wastewaters is a direct reflect of the rapid spreading of UK variant in Paris since December 2020. Observed proportions are close to those found by two recent intensive sequencing studies in French patients (green dots in figure 2).

If the spreading of the B.1.1.7 variant was relatively slow during January 2021, the dynamic of its diffusion into the Parisian population has accelerated significantly since January 27th. It is to note that the relative proportion of del69-70 carrying variants in raw wastewater was quite close to the estimation of B.1.1.7 variant in positive patients confirmed by virus sequencing. More Information about shedding of virus from B.1.1.7 infected patients need to be collected to compare with 2020 variants excretion in faeces. A recent publication showed that a longer shedding time in the upper respiratory tract could occur in B.1.1.7 infected patients (Kissler et al. 2021). If this would be confirmed for enteric shedding, these results could influence the ratio del69-70/wild-type in wastewater.

To our knowledge, the sequencing of viral RNA isolated from patients was not routinely done up to the end of January 2021 and only few sequencing studies were occasionally performed in order to evaluate the progression of 2021 variants in France. Sequencing results have shown differences in mutation proportions between different countries/regions, but in all cases, SARS-CoV-2 strains containing common nucleotide sequences at different positions were selected (Martin et al. 2020). Theses mutations could be more simply detected by RT-qPCR. RT-qPCR tools specifically targeting relevant polymorphisms (del69-70, N501Y or E484K) in raw wastewater provide a faster and cheaper tool for evaluating spread of variants in populations once relevant mutations have been identified, compared to sequencing approaches. This adds a new facet to the macro-epidemiological indicator that is the analysis of wastewater by combining the frequency of sampling already in place with the specificity of this search for specific mutations. Moreover our group already showed that the integrity of SARS-CoV-2 is significantly impacted during its transportation from stool to wastewater (Wurtzer, Waldman, et al. 2020). If the viral genome is fragmented, the identification of variant based on the detection of mutation combinations on the same strand by sequencing could be illusory.

Based on this proof-of-concept, various RT-qPCR test with high sensitivity and specificity need to be developed for wastewater matrix, with priorizing the polymorphisms N501Y and E484K.

## Data Availability

all data are available

